# Discrimination of Vocal Folds Lesions by Multiclass Classification using Autofluorescence Spectroscopy

**DOI:** 10.1101/2023.05.11.23289778

**Authors:** Olivier Gaiffe, Joackim Mahdjoub, Emmanuel Ramasso, Olivier Mauvais, Thomas Lihoreau, Lionel Pazart, Bruno Wacogne, Laurent Tavernier

## Abstract

**Objectives:** The diagnosis of vocal fold cancer currently relies on invasive surgical biopsies, which can compromise laryngeal function. Distinguishing between different types of laryngeal lesions without invasive tissue sampling is therefore crucial. Autofluorescence spectroscopy (AFS) has proved to be efficient as a non-invasive detection technique but has yet to be fully exploited in the context of a multi-class tissue analysis. This study evaluates whether AFS can be used to discriminate between different types of laryngeal lesions in view of assisting in vocal fold surgery and preoperative investigations.

**Materials and methods:** *Ex vivo* spectral autofluorescence scans were recorded for each sample using a 405-nm laser excitation. A total of 1308 spectra were recorded from 29 vocal fold samples obtained from 23 patients. Multiclass analysis was conducted on the spectral data, classifying lesions either as normal, benign, dysplastic, or carcinoma. The results were compared to histopathological diagnosis.

**Results:** Through an appropriate selection of spectral components and a cascading classification approach based on artificial neural networks (ANN), a classification rate of 97% was achieved for each lesion class, compared to 52% using autofluorescence intensity.

**Conclusion:** The study demonstrates the effectiveness of AFS combined with multivariate analysis for accurate classification of vocal fold lesions. Comprehensive spectral data analysis significantly improves classification accuracy, even in challenging situations such as distinguishing between malignant and premalignant or benign lesions. This method could provide a way to perform *in situ* mapping of tissue states for minimally-invasive biopsy and surgical resection margins.

## Introduction

Early detection is a major issue in the management of laryngeal cancer, as delay can lead to a poor prognosis and or the necessity of aggressive treatments with potential severe functional impairment. Currently, histopathology remains the gold standard for the identification and extension assessment of the lesions. This analysis involves collecting biopsies, generally during microlaryngoscopy under general anesthesia, making it an inherently invasive method. However, microscopic observation alone cannot always assess cancerous or premalignant lesions and their exact delineation. Moreover, the need to obtain clear surgical margins and the risk of false negatives due to incorrect sample areas can lead to an increased number of biopsies. In the case of vocal folds, repeated biopsies can lead to severe voice and swallowing disorders.

Therefore, finding minimally invasive diagnosis techniques is a major challenge. Many complementary diagnostic tools have been developed to help clinicians^1^. Because of their non-invasive nature, optical techniques are among the most promising methods^2,3^. Current data^4,5^ from the literature support the idea that many cellular and subcellular changes can be detected using these methods.

Autofluorescence endoscopy imaging (AFEI) and autofluorescence spectroscopy (AFS) have been gaining interest during the last decades. They are accurate and fast optical methods that can be used early^6,7^ and allow real-time detection of malignancies^8,9^. These techniques are based on the measurement of tissue fluorescence emission. Under ultraviolet or blue irradiation, several endogenous fluorophores such as collagen, elastin, NADH, flavin, or porphyrins produce radiation in the visible wavelength region. The shape of the fluorescence spectra depends on the excitation wavelength, on the presence of fluorophores and on the optical properties of tissues. Typically, molecular changes in carcinogenesis lead to distinct fluorescence spectra^10^.

In the field of laryngeal oncology, many studies focused on using the intensity of the fluorescence signal (*Ig*) of the tissues as a biomarker to discriminate lesions^11–13^. Results were encouraging but showed poor specificity, as many benign pathologies caused a decrease in fluorescence intensity. A small number of studies focused on fluorescence spectroscopy^10,14–16^ and showed differences between spectral patterns of normal and malignant tissues of the larynx. However, discriminating benign from cancerous lesions, or premalignant from malignant lesions remained impossible in these studies. Another difficulty was the way of selecting significant information contained in the spectral data. Altogether, the diagnostic benefit of AFS was low. Yet, AFEI and AFS have been successfully applied to cancer detection in a number of other organs, notably in the oral cavity^16,17^. Indeed, *ex vivo* and *in vivo* studies have shown the effectiveness of AFEI and AFS for the diagnosis of oral squamous cell carcinoma with sensitivity 0.82-1.00 and specificity 0.63-1.00 depending on the series^18–21^.

The objective of the present study was therefore to assess the accuracy of AFS in distinguishing between different types of vocal cord lesions. Our approach involved selecting a sufficient number of spectral components and employing a cascading classification method based on artificial neural networks (ANN) to achieve multi-class discrimination. To evaluate the effectiveness of the method, classification models based on global fluorescence intensity or using discriminant analysis were used for comparison.

## Materials and methods

### Clinical Study Design

In this prospective study, we examined patients undergoing vocal folds surgery at the Besançon University Hospital between 2014 and 2016. “Fluorocord” trial received a favorable opinion from French ethical committee (CPP Est-II) on June 2014 (NCT03585075). Informed written consent was obtained from each patient. Biopsy sites for lesions were selected on a visual examination basis by the surgeon. Normal vocal fold biopsies were taken from total laryngectomy surgical specimens or from cordectomy specimens for non-tumoral indication. After AFS recording, biopsies were embedded in 10% neutral formalin and sent to histopathology for diagnosis confirmation.

### Fluorescence Spectroscopy Instrumentation

The point-scan method was used to obtain a hyperspectral fluorescence image of 1.4 by 1.4 mm^2^ with a pitch of 200 μm of each biopsy. The excitation wavelength set at 405 nm (Flexpoint) was carried by a fiber-optic probe (Avantes) consisting of seven 200-μm-core fibers, one used for excitation and the others for collection. A bandpass filter centered at 405 nm (±20 nm) was placed on the excitation path and a low pass filter at 437 nm was used to remove the excitation light from the collection path. The fluorescence spectrum was then measured by a spectrometer (Ocean Optics USB 4000ES). Biopsies were placed on a specific sample holder and kept moist with saline solution. A 170-μm thick cover glass was placed atop the biopsy to ensure sample flatness. The probe was located 100 μm away from the cover glass. Each measurement point was associated with a fluorescence spectrum recorded in the wavelength range 450-750 nm sampled by 1550 measurement points spaced 0.20 nm apart. Spectra saturating the sensor or having a signal-to-noise ratio < 10 dB were removed from the study.

### Features Extraction

The spectral features were estimated using a Principal Component Analysis (PCA) in order to reduce the high data dimensionality^22^. The result of PCA is a set of new dimensions called Principal Components (PCs) which are uncorrelated and are computed by preserving as much variance as possible. In the present case, we selected a number of PCs allowing us to obtain at least 99.9% of the amount of variance contained in the original data set. The fluorescent intensity (*Ig*) was also extracted from fluorescence spectra in order to be used as a reference during the classification step. This descriptor (*Ig*) was estimated by calculating the integral of the spectrum between 437 - 750 nm.

### Tissue classification methods

Histopathologic diagnosis was performed by grouping all the biopsies into four categories: normal, benign, dysplasia, and carcinoma. Histopathologic analyses were realized by an independent pathologist.

We chose to cascade three binary models to achieve classification into the 4 categories (**Fig 1**). Each model or sub-model extracts one class and remaining classes become the input of the next sub-model until all classes are separated. The interest of this cascading classification method has been proven for Raman spectroscopy-based classification of tumor and non-tumor tissues^23^. We chose to isolate normal tissue first, then benign tissue and finally separate precancerous from cancerous. The final classification of a biopsy into one of the four aforementioned categories was performed by taking the predominant category (majority vote) of its spectra.

**Figure 1.**
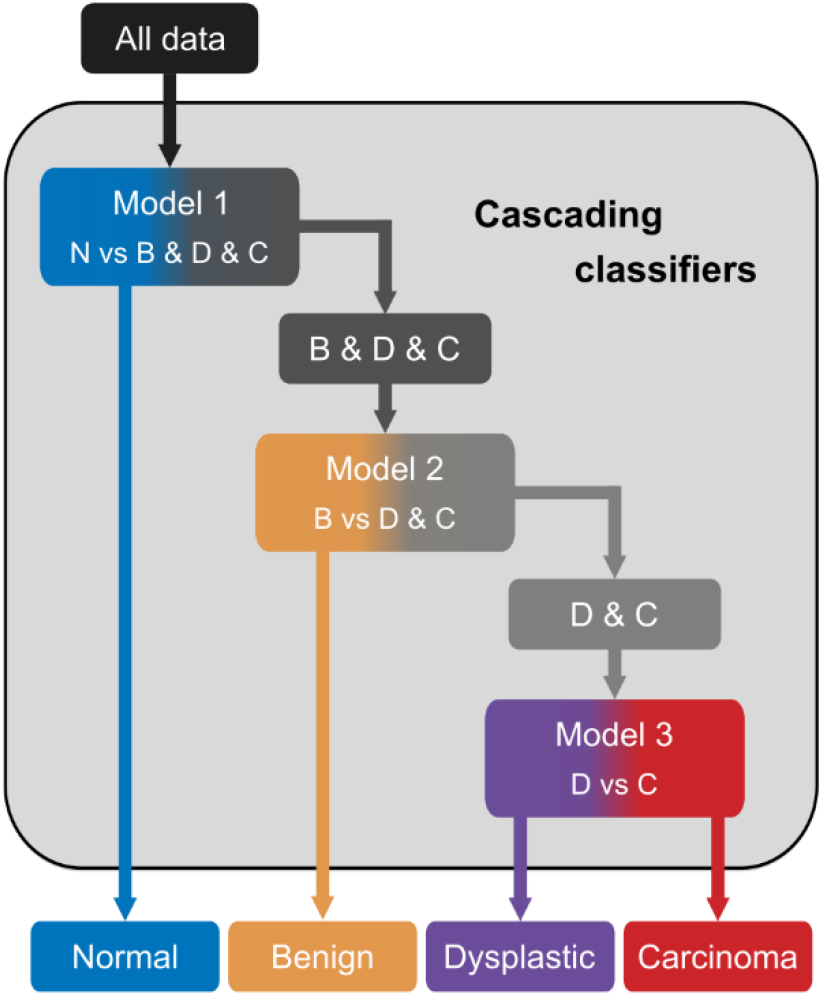
Cascading Classification Method. Cascading Classification using three different models to separate 4 groups: normal (N), benign (B), dysplastic (D), and carcinoma (C).

Two classification methods^24^, Discriminant Analysis (DA) and ANN were used and compared. DA can separate classes by considering either a linear (LDA) or a quadratic (QDA) combination of features. ANN is known to be a universal approximator which can be used when the data generating process is unknown or complex. The ANN used in the present study consists of a single hidden layer with 10 neurons. The methods implemented were PCA+DA and PCA+ANN when analyzing spectral features. A 5-fold cross-validation was used to validate the robustness of each classification model and method.

From the classification results, we evaluated the diagnostic accuracy (Acc), the sensitivity (Se), the specificity (Sp) and the Youden index^25^ (J) which is calculated as J = Se+Sp-1, thus giving an optimal value of the discrimination threshold. Clopper-Pearson 95% confidence intervals were calculated for Se, Sp, and Acc. All data processing and statistical data analysis were performed using MATLAB™ 2019b (MathWorks Inc., Natick MA).

## Results

### Cohort Characteristics

A sample of 29 biopsies were used from 23 patients. For 6 patients, 2 biopsies were collected on each vocal cord. According to the histological analysis, 5 biopsies were classified as dysplasia (including 2 moderate to severe dysplasia and 3 carcinomas *in situ* (CIS)), 10 as carcinoma (including 8 Squamous Cell Carcinoma (SCC) and 2 spindle cell carcinomas), 5 as normal and 8 as benign (including 4 polyps, 2 inflammatory lesions, 1 ulceration with granuloma, 1 papilloma and 1 leukokeratosis) (**Table 1**). A total of 1308 spectra was recorded and divided as follows: 245 normal spectra, 409 benign, 227 dysplastic, and 427 malignant.

**Table 1:** Cohort Characteristics.

| Patient | Sex | Biopsy (n°) | Group | Histology |
| --- | --- | --- | --- | --- |
| A | M | 1 | N | Normal |
| B | F | 2 | C | SCC |
| C | F | 3 | D | Moderate to severe dysplasia |
|  |  | 4 | C | SCC |
| D | F | 5 | B | Myxoid polyp |
| E | M | 6 | C | SCC |
| F | M | 7 | C | SCC |
| G | M | 8 | D | CIS |
| H | M | 9 | B | Polyp |
| I | M | 10 | C | SCC |
|  |  | 11 | D | Moderate dysplasia |
| J | M | 12 | D | CIS |
|  |  | 13 | N | Normal |
| K | M | 14 | C | SCC |
|  |  | 15 | B | Inflammation |
| L | M | 16 | B | Mucosal ulceration with granulation tissue |
| M | M | 17 | B | Inflammatory polyp |
| N | M | 18 | B | Leukokeratosis |
| O | M | 19 | B | Inflammation, hyperplasia |
| P | M | 20 | C | Spindle cell carcinoma |
| Q | M | 21 | D | Severe dysplasia to CIS |
| R | M | 22 | C | Spindle cell carcinoma |
|  |  | 23 | N | Normal |
| S | M | 24 | B | Papilloma |
| T | M | 25 | C | SCC |
| U | M | 26 | C | SCC |
| V | M | 27 | B | Pseudo-polyp |
| W | M | 28 | N | Normal |
|  |  | 29 | N | Normal |

Abbreviations: Male (M), Female (F), Normal (N), Benign (B), Dysplastic (D), Carcinoma (C), Squamous Cell Carcinoma (SCC), Carcinoma *In Situ* (CIS).

### Spectral Response Patterns

The intensity Ig for each of the recorded data is represented in the box plot reported in **Figure 2a**. The fluorescence intensity decreases from the normal to the benign group. The median value of the benign group is close to the carcinoma group. Normalized area-under-the-curve (nAUC) spectra, typical of each lesion group, are displayed in **Figure 2b**. In this example, the benign case corresponded to an inflammatory polyp, and a noticeable shift in the central wavelength was observed. The dysplastic case was diagnosed as CIS. Interestingly, the corresponding spectrum did not exhibit any discernible differences compared to the normal case. On the other hand, the spectrum obtained from an invasive squamous cell carcinoma showed a distinct and highly differentiated spectral shape, characterized by a prominent peak at 632 nm. However, it is important to note that this specific spectral characteristic, although easily identifiable, was not consistently observed in all the cancerous biopsies included in our study.

**Figure 2.**
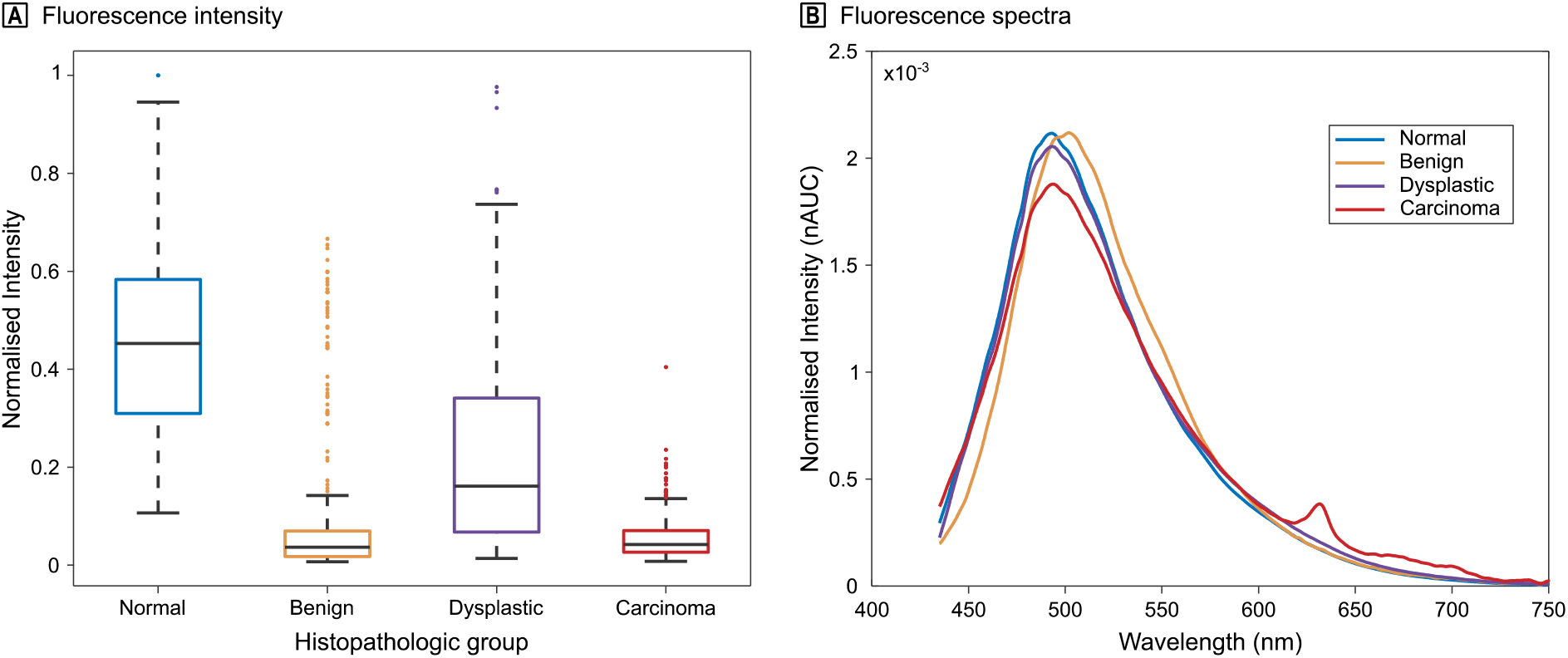
Fluorescence Intensity and Spectral Dataset. A, fluorescence intensity Ig of the four categories for all measurements normalized by the maximum of the dataset. The box boundaries indicate the first and third quartile and the black line indicates the median. The upper and lower whiskers are specified as 1.5 times the interquartile range. Dots denote outliers. B, normalized area-under-the-curve (nAUC) spectra representative of the 4 groups: normal (biopsy 1), benign (biospy 17), dysplastic (biospy 12), carcinoma (biopsy 14).

### Classification of sub-model

Initially, each of the three sub-models was studied individually to determine the optimum classification parameters, i.e. the number of PCs to be considered as well as the type of classification method to be used.

The objective of the first sub-model was to distinguish healthy tissue from diseased tissue. As shown in **Table 2**, the classification results based on Ig intensity were equivalent regardless of the classification method used. Se, Sp, and Acc ranged from 0.87-0.89, 0.80-0.84, and 0.87, respectively. J-index was used to determine the optimal value of the classification results. J was slightly higher for ANN compared to DA with values of 0.71 and 0.69 respectively. Classification results obtained from spectral data were superior to the results obtained using Ig. **Figure 3a** shows the evolution of the J-index as a function of the number of PCs used for DA and ANN. We observed that ANN remains higher than DA for all number of PCs used. In the case of ANN, the J-index rapidly increased up to 5 PCs and then reached a plateau, contrarily to the classification by DA which required 8 PCs, and therefore more information, to stabilize. The fluorescence spectrum provided a significant improvement leading to an increase in the J-index of 12% (DA) and 13% (ANN). The maximum value was 0.97 (CI 0.97-1.00), obtained using ANN with a sensitivity of 0.99 (CI 0.99-1.00) and a specificity of 0.98 (CI 0.95-0.99).

**Table 2:** Classification results of sub-models.

|  |  | Mean (CI 95%) |  |  |  |
| --- | --- | --- | --- | --- | --- |
| Classifier |  | Sensitivity | Specificity | Accuracy | J-index |
| N vs (B, D & C) “sub-model 1” |  |  |  |  |  |
| Intensity | DA | 0.89 (0.87-0.91) | 0.80 (0.74-0.85) | 0.87 (0.85-0.89) | 0.69 (0.61-0.76) |
|  | ANN | 0.87 (0.85-0.89) | 0.84 (0.79-0.88) | 0.86 (0.84-0.88) | 0.71 (0.64-0.77) |
| Spectra | DA | 0.92 (0.90-0.94) | 1.00 (0.99-1.00) | 0.94 (0.92-0.95) | 0.92 (0.89-0.94) |
|  | ANN | 0.99 (0.98-0.99) | 0.98 (0.95-0.99) | 0.99 (0.98-0.99) | 0.97 (0.93-0.98) |
| B vs (D & C) “sub-model 2” |  |  |  |  |  |
| Intensity | DA | 0.56 (0.52-0.60) | 0.86 (0.82-0.89) | 0.68 (0.65-0.70) | 0.42 (0.34-0.39) |
|  | ANN | 0.85 (0.82-0.88) | 0.44 (0.39-0.49) | 0.69 (0.66-0.72) | 0.29 (0.21-0.37) |
| Spectra | DA | 0.60 (0.56-0.64) | 0.99 (0.98-1.00) | 0.75 (0.72-0.78) | 0.59 (0.54-0.64) |
|  | ANN | 0.86 (0.83-0.89) | 0.86 (0.82-0.89) | 0.86 (0.84-0.88) | 0.72 (0.65-0.78) |
| D vs C “sub-model 3” |  |  |  |  |  |
| Intensity | DA | 0.96 (0.94-0.98) | 0.53 (0.46-0.60) | 0.81 (0.78-0.84) | 0.49 (0.40-0.58) |
|  | ANN | 0.95 (0.93-0.97) | 0.55 (0.48-0.62) | 0.81 (0.78-0.84) | 0.50 (0.41-0.59) |
| Spectra | DA | 0.95 (0.93-0.97) | 0.78 (0.72-0.83) | 0.89 (0.87-0.91) | 0.73 (0.65-0.80) |
|  | ANN | 0.97 (0.95-0.98) | 0.92 (0.85-0.95) | 0.95 (0.93-0.97) | 0.89 (0.83-0.93) |

Abbreviations: Normal (N), Benign (B), Dysplastic (D), Carcinoma (C), Discriminant Analysis (DA), Artificial Neural Network (ANN), Accuracy (Acc), Sensitivity (Se), Specificity (Sp), Youden index (J).

**Figure 3.**
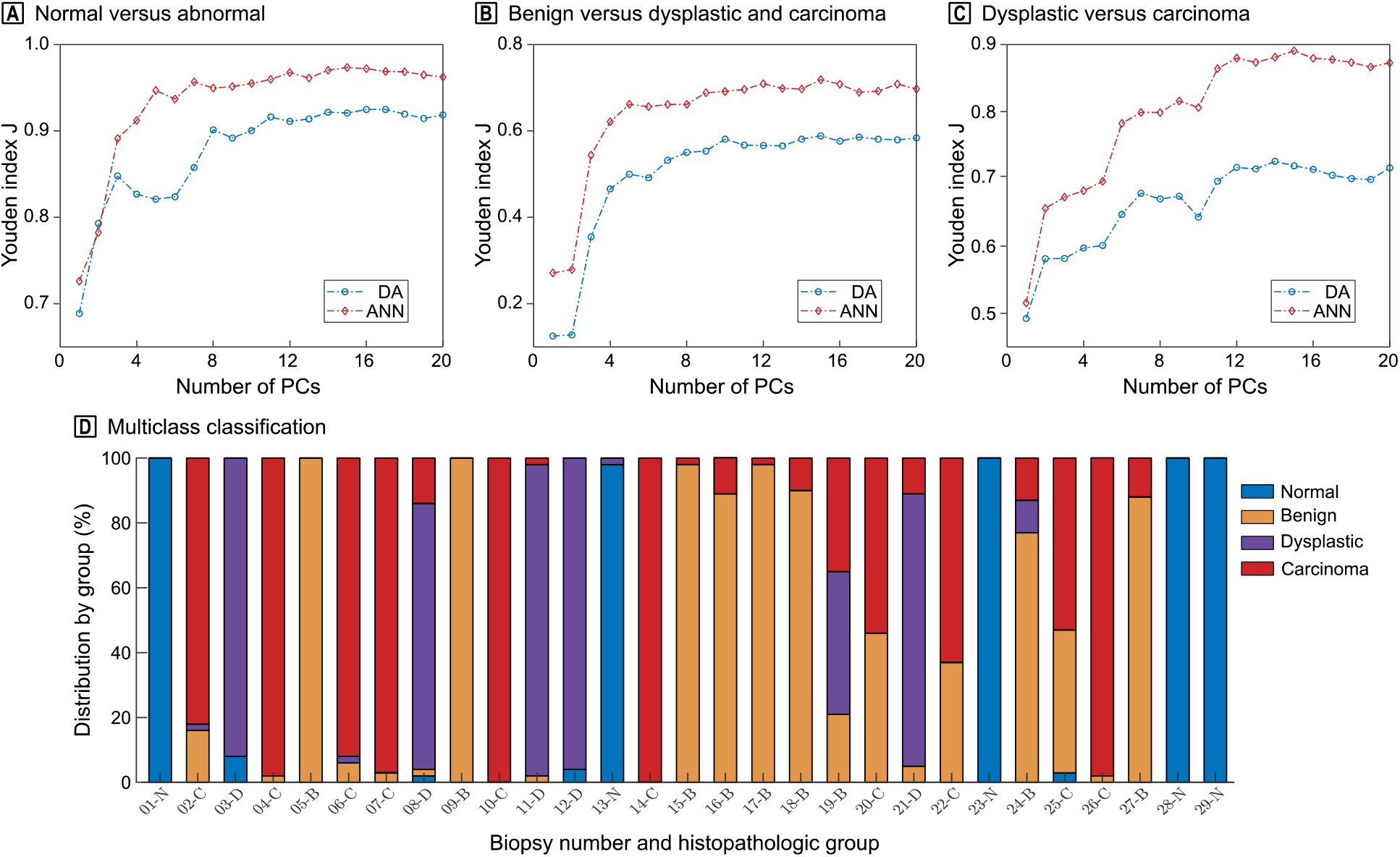
Classification Results. A, B and C, Youden index (J) according to the number of principal components for DA and ANN classifiers. D, multiclass classification results by cascading classifiers represented by the percentage of membership per class of each biopsy.

The second sub-model aimed at separating benign tissue from precancerous and cancerous tissue. The results obtained using Ig (**Table 2**) featured Se and Sp values near 0.50. DA gave the largest J-index with a value of 0.42 (CI 0.34-0.49) which was 14% higher compared to ANN. The estimated Se and Sp were 0.56 (CI 0.52-0.60) and 0.86 (CI 0.82-0.89). The addition of spectral information allowed, in this case as well, to increase the classification performance. The trend was similar to first model with a rapid increase up to 5 PCs followed by a plateau (**Fig 2b**). For all PCs, the results obtained by ANN were superior to DA. The maximum value of J-index was 0.72 (CI 0.65-0.78). The corresponding Se and Sp parameters were 0.86 (CI 0.83-0.89) and 0.86 (CI 0.82-0.89), respectively. The fluorescence spectral information made it possible to obtain a classification gain on J-index of 15% compared to intensity.

The last sub-model sought to discriminate dysplastic tissue from malignant tissue. The classification results using Ig were similar regardless of the classification method (**Table 2**). The J-index was between 0.49 and 0.50 for Se and Sp varying from 0.94 to 0.96 and from 0.53 to 0.55, respectively. As in the previous cases, using spectral information increased the classification efficiency by 20% compared with Ig. The increase in the J-index (**Fig 2c**) with the number of PCs showed three distinct plateaus. The model required a larger number of PCs and therefore more spectral information (11 PCs) to reach the last plateau. The J-index by ANN was higher with a value of 0.89 (IC 0.83-0.93) against 0.73 (IC 0.65-0.80) by DA. The Se and Sp parameters were 0.97 (CI 0.95-0.98) and 0.92 (CI 0.88-0.95), respectively.

### Multiclass Classification

An overall classification model was eventually implemented following the cascading classification method shown in **Fig 1**. Taking each spectrum as independent, the accuracy of the multiclass model was 89% with 57% of the errors related specifically to 4 biopsies diagnosed as inflammation - hyperplasia, SCC and two spindle-cell carcinomas. The misclassification between the carcinoma and benign classes alone accounts for 66% of the errors.

For each biopsy, a membership percentage by class was obtained (**Fig 2d**). Considering a single class per biopsy, defined by the predominant category contained in the analyzed scan, leads to a result of 28 out of 29 well-classified biopsies, i.e. to an accuracy of 0.97 versus 0.52 with fluorescence intensity alone.

## Discussion

In this prospective trial, we demonstrated that autofluorescence spectroscopy is an efficient way of discriminating vocal folds lesions into 4 classes. The use of all the information contained in fluorescence spectra combined with multivariate analysis provided good classification performance, even in situations where distinction could not be made in the literature^15^ (e.g. malignant vs. premalignant or benign lesions). Overall, 28 out of the 29 lesions were correctly recognized by our multiclass model.

In order to compare our classification results based on the analysis of fluorescence intensity (Ig) only, we extracted the Youden index from data taken from the literature^11,13^. Youden indices ranging from 0.76 to 0.90 were reported, higher than the 0.71 obtained in the present case. Indeed, in these studies, the authors used the AFEI to select the area to be biopsied. The clinicians therefore had additional information such as the structural modifications of the lesions, the tissue architecture or the neovascularization.

Only 3 studies evaluated AFS on laryngeal pathologies to separate healthy tissue from diseased tissue^10,15,16^, yet excluding benign lesions from the analyses. The conclusions were that AFS could not, or barely, improve the diagnosis compared to AFEI. Our hypothesis is that the recorded spectral data were not fully exploited in these works. Arens *et al*. only examined average spectra maxima, which actually amounts to exploiting Ig, while Rydell *et al*. compared intensity ratios at different wavelengths. Only Eker *et al*. used several multivariate analysis methods, including PCA with 4 PCs (4 PCs + QDA), and found a J-index of 0.81. By applying the same restrictions to our classification parameters, we obtained a similar result of 0.82. Increasing the J-index actually required analyzing more PCs: comparatively, we needed at least 8 PCs to obtain 0.90 with QDA and at least 7 PCs with ANN to obtain a J greater than 0.95.

Regarding multivariate analyses, PCA+ANN proved to be more effective than PCA+DA, particularly in isolating benign cases and in separating dysplasia from carcinoma. These results support previous studies in the field of oral oncology: Van Staveren *et al*. obtained Se of 0.86 and Sp of 1.00 when isolating leukoplakia with ANN^26^. Nayak *et al*. also found excellent diagnostic performances using AFS to discriminate between normal, precancerous and malignant lesions of the oral cavity (excluding benign lesions), with a superiority of ANN^8^.

In our study, the model 2 isolating the benign cases had the weakest classification result. The inclusion of benign samples leads to classification difficulties, as fluorescence characteristics are very similar to those of malignant lesions^27,28^. This may be due to excitation light absorption by hemoglobin and inflammatory cells^17,29^. Another difficulty was observed in the literature to discriminate between dysplastic and malignant lesions either with AFEI^30^ or with AFS^15^. In our model 3, a distinction can be clearly made using PCA+ANN with 15 PCs. To our knowledge, our model is the first to successfully discriminate between dysplasia or CIS and invasive cancer for vocal cord tissue. This supports the idea that using more spectral information leads to better classification results.

Concerning the multiclass classification, a single biopsy was incorrectly classified as dysplastic when it was defined as benign by the pathologist. The classification result of the sample was dispersed into 3 classes, with 21% of information classified as benign, 44% as dysplastic, and 35% as carcinoma. the histology of the sample concluded with a very hyperkeratotic inflammatory tissue. This is consistent with previous studies^27,29^ that found SCC false negatives in AFEI related to extreme hyperkeratosis. Indeed, hyperkeratosis leads to poor penetration of light irradiation, therefore reducing fluorescence emission from the underlying cell layers and thus mimicking a dysplastic or a malignant lesion. Moreover, this patient had many local recurrences during his follow-up and a verrucous carcinoma was eventually diagnosed. This entity is a rare and low-grade variant of SCC, which frequently leads to diagnosis difficulties.

Three correctly classified carcinoma samples showed between 37% and 46% of their distributed spectra as benign. Two of them were spindle-cell SCC: rare, aggressive, and poorly differentiated variants of SCC which are often diagnostic challenges for pathologists. Studies showed that this type of tumor could often mimic benign mesenchymal proliferation^31,32^. We hypothesize that these histopathologic features may influence fluorescence findings as well. The third case was a micro-invasive SCC with underlying inflammatory stroma, which may account for our results.

The main limit of our study was the number of samples per class. The monocentric nature of the study also limits its generalization at this stage. A multicenter trial would therefore allow increasing the number of samples and expanding the population. Eventually, this *ex vivo* study only partially represents the clinical reality as it limits the presence of blood that could disturb the sample measurements in an actual surgical context. It will thus be necessary to confirm this model *in-vivo*.

## Conclusion

Autofluorescence spectroscopy combined with multivariate analysis has proven effective in classifying vocal cord lesions. AFS was shown to increase classification accuracy into 4 major categories up to 97%, versus 52% when considering the fluorescence intensity alone. A comprehensive analysis of the principal components allowed distinguishing lesion types, even in difficult situations. In particular, discriminating malignant from precancerous or benign lesions was successfully achieved. In the future, AFS could be implemented as an accurate minimally-invasive diagnostic tool, next to clinical examination, in the management of laryngeal pathology. Also, the multispectral mapping process could be an aid for surgical resection margins of malignant lesions.

## Data Availability

All data produced in the present study are available upon reasonable request to the authors

## Article information

### Additional Contributions

We thank the patients for their participation.

### Additional Information

Research data are stored in an institutional repository and will be shared upon request to the corresponding author.

